# Rare variants of the glucagon-like peptide-1 receptor (GLP1R) gene are overrepresented in a severe obesity cohort and associated with type 2 diabetes in the UK Biobank

**DOI:** 10.1101/2023.05.22.23290347

**Authors:** Dale Handley, Sumaya Almansoori, Mitra S. Sato, Hasnat Amin, Suzanne Alsters, Harvinder Chahal, Sanjay Purkayastha, Kevin G. Murphy, Mieke van Haelst, Carel W le Roux, Tricia Tan, Robin G Walters, Fotios Drenos, Alexandra I Blakemore

**Affiliations:** College of Health and Life Sciences, Brunel University London, Uxbridge, UK; Mohammed bin Rashid University of Medicine and Health Sciences, Dubai, United Arab Emirate; Department of metabolism, Digestion, and Reproduction, Imperial College London, London, UK; South West Thames Regional Genetics Service, St George’s University Hospitals NHS Foundation Trust, London, UK; Imperial College Healthcare NHS trust, St. Mary’s hospital, London, UK; Section of Clinical Genetics, Department of Human Genetics, Amsterdam Reproduction and Development, Amsterdam University Medical Centers, Amsterdam, The Netherlands; Diabetes complications research centre, Conway institute, University College Dublin, Dublin, Republic of Ireland; MRC Population Health Research Unit, University of Oxford, Oxford, UK; College of Medicine, Nursing, and Health science, University of Galway, Galway, Republic of Ireland

## Abstract

**Introduction:** Glucagon-like peptide 1 (GLP1) agonists are highly effective agents for the treatment of obesity and type 2 diabetes (T2D). GLP-1 is also implicated in outcomes of bariatric surgery, including appetite changes and T2D remission. Rare, potentially deleterious mutations in the glucagon-like peptide 1 receptor gene (*GLP1R*) may, therefore, have important implications for pathogenesis of obesity and T2D, and for response to therapeutic interventions.

**Methods:** A custom Axion genotyping array, including 117 rare predicted-deleterious *GLP1R* mutations (MAF<0.01 in gnomAD, CADD-PHRED m >= 15), was used to screen 1714 unrelated adults with BMI >35 kg/m^2^ from the PMMO study. We also examined the UK Biobank (UKB) exome sequence dataset for rare, predicted-deleterious *GLP1R* variants and tested their effects on weight and glycaemia-related traits.

**Results:** Thirty-four PMMO participants carried one of the 117 *GLP1R* variants screened (11 might have been expected using the sum of their gnomAD control MAFs). These 8 variants were associated with T2D in the UKB and subsequent gene-level analysis of the UKB exome sequence dataset (629/39,274 carriers) confirmed that rare *GLP1R* variants are associated with increased risk of T2D (OR=1.58), as well as with higher HbA1c levels (p= 0.039). Furthermore, our data highlight a potential interaction of these variants with body mass index.

**Conclusion:** Rare, potentially deleterious *GLP1R* mutations is associated with increased T2D risk, as well as higher HbA1c in UKB participants without diabetes. Future studies should examine the implications of *GLP1R* mutations for response to GLP1 agonist treatment and explore the observed interactions with obesity in T2D risk, including in larger cohorts with obesity.

## Introduction

GLP1 agonists are well-established treatments for type 2 diabetes (T2D) and obesity (Kelly *et al*., 2013; Hinnen, 2017; Webb *et al*., 2017; Patel and Smith, 2021; Wilding *et al*.,2021). Semaglutide results in weight loss outcomes almost comparable to bariatric surgery (Wilding *et al*., 2021). Furthermore, more recently developed agents that combine GLP1 and GIP activity (eg. Tirzepatide) or GLP1 with amylin activity result in even more weight loss (Enebo *et al*., 2021; Jastreboff *et al*., 2022; (D’ascanio *et al*., 2023).

GLP1 acts on the GLP1 receptor (GLP1R) to drive downstream effects, including increased insulin release, insulin sensitivity and diminished appetite (Bloemendall *et al*., 2014; Sandoval *et* al., 2015; Jiang *et al*., 2018). GLP1 is released post-prandially by intestinal L-cells (Lim and Brubaker, 2009), and binds to the glucagon-like peptide receptor (GLP1R) increasing satiety, increasing insulin secretion (Meloni *at al*., 2013; Jones *et al*., 2018), and delaying gastric emptying and gut motility (Marathe *et al*., 2013; Maselli and Camilleri, 2021), alongside other effects (as reviewed by Drucker, 2018).

GLP1 is also implicated in the decreased appetite and improved glycaemic control following gastric bypass or sleeve gastrectomy surgeries (as reviewed by Hutch and Sandoval, 2017) Bariatric surgery increases post-prandial GLP1 secretion (Jiranpinyo *et al*., 2018). Furthermore, antagonism of GLP1R following bariatric surgery in mice decreases weight loss and increases weight regain (Larraufie *et al*., 2019). The effects of bariatric surgery on T2D are particularly striking: many individuals experiencing increased insulin sensitivity and improved glycaemic control long before clinically significant weight reduction occurs (as reviewed by Laferrere *et al*., 2018). This often includes remission of T2D. Durability of this effect and the importance of GLP1 in long-term outcomes of bariatric surgery are well characterised (Werling *et al*., 2014; Amouyal and Andreelli, 2016; Papamargaritis and le Roux, 2021), Thus, the efficacy of GLP1 agonists and bariatric surgery for the treatment of T2D and obesity may be dependent on a well-functioning GLP1 receptor. Several common genetic variants have been identified as being detrimental to the response to GLP1R agonists (Rathmann and Bongaerts, 2021), but the prevalence of rare and potentially deleterious *GLP1R* variants and their effects on BMI, T2D risk, and response to therapeutic interventions are yet to be determined. Here, we describe initial analysis of rare *GLP1R* variants in a clinical cohort of unrelated adults with BMI >35kg/m^2^ and providing further evidence for the role of GLP1R variants in T2D risk in the UK Biobank (UKB).

## Methods

### Ethics statement

Ethical permission for the sample collection, genetic analysis, and use of data from the Personalised Medicine for Morbid Obesity study was approved by the NRES Committee London Riverside (REC reference 11\LO\0935), and the research was performed in accordance with the principles of the Declaration of Helsinki. All participants gave informed consent. Permission for use of the UKB for the investigation of obesity-related traits is provided under UKB application number 62265. This research was also approved by Brunel University London Ethics committee application number 26158.

### Cohort descriptions and data collection

#### Personalised Medicine for Morbid Obesity

The Personalised Medicine for Morbid Obesity (PMMO) NHS-registered portfolio trial (NCT01365416; https://clinicaltrials.gov/ct2/show/NCT01365416) includes a total of 2556 unrelated adult individuals, recruited between 2011 and 2021, BMI>35 kg/m^2^ with significant co-morbidities). Participants were recruited either prospectively across five different hospitals in the United Kingdom (Chelsea and Westminster Hospital, London; The Imperial College NHS Weight Centre, St. Mary’s Hospital, London; Royal Derbyshire Hospital, Derby; Charing Cross Hospital, London; Sunderland Royal Hospital, Sunderland) or retrospectively using NHS medical records from the Imperial Weight Centre between 2012 - 2021. For those recruited before surgery, the data used here were collected at three time points: pre-surgery (as close to the date of bariatric surgery as possible, or at recruitment for those who did not undergo surgery), 1-year after surgery, and 2-years after surgery. For those recruited after surgery, retrospective data was collected from NHS electronic records for the timepoint closest to the recorded surgery date (if available), and as close as possible to 1-year after surgery, and 2-years after surgery. Data with individuals who were lost to follow up were retained where excessive (>50%) of baseline data was not available. For both sets of data, T2D status at baseline was defined as any previous clinical diagnosis of T2D.

#### UK biobank

The UKB is a prospective cohort study including >500,000 individuals of predominantly White British ancestry, aged 40-69 years, examined across 22 assessment centres in England, Wales, and Scotland, with the primary data collection occurring between years 2006-2010 (Sudlow *et al*., 2015). The median age of participants in the UKB at recruitment was 56.5, and self-reported females represented 54.4% of the cohort. To date, information is available for over 7,000 measures, derived from questionnaires, cognition tests, imaging, medical records, and biochemical analysis of blood samples.

### Genotyping of samples and quality control

#### PMMO

Genomic DNA was prepared from either blood or saliva samples by standard methods. In total, 1766 samples from the PMMO cohort underwent genotyping using a custom Axiom genotyping array prepared by ThermoFisher, as previously described (ThermoFisher, 2021). Genotyping was performed according to ThermoFisher guidelines (ThermoFisher, 2021) by the Oxford Genomics Centre (Oxford, UK) and processed using a GeneTitan Multi-channel instrument. Probe clustering was performed using the AxiomGT1 algorithm from the Axiom analysis suite V5.1.1 (ThermoFisher, 2021). The rare heterozygosity adjustment available within the Axiom software was used to correct for multi-probe mismatches, which substantially improves correct rare variant calls for very rare variants (MAF > 1×10^−4^, Hsun-sun *et al*., 2021). Initial genotyping filtering was conducted according to the Axiom array guidelines, with the dish quality control threshold, which determines the presence of genotyping plate-specific effects, set to 82% (Axiom, 2021). In total, 117 rare deleterious *GLP1R* variants were included, based on combined annotate dependent depletion normalised score (CADD PHRED) threshold score >=20, rarity (MAF < 0.01 in the overall gnomAD population) (Karczewski *et al*., 2020). All *GLP1R* rare variants included in the genotyping array are shown in supplementary table 1. Samples that did not pass the QC metrics provided above (n=25), that were duplicates (n = 22), showed a high degree of heterozygosity or relatedness (n = 0), or which displayed discordant genetic and self-reported sex (n = 5) were removed from further analysis. Genetic variants in *GLP1R* with a low call rate (< 93% call rate) would also have been removed from further analysis (n = 0). In total, 1714 samples remained following sample QC, and all 117 rare, deleterious *GLP1R* variants survived the variant QC. The cluster plots for all *GLP1R* variants included in this analysis were inspected visually using the Axiom analysis suite to ensure a sufficiently clear separation between the clusters.

### UK biobank whole exome sequencing

For the 50k whole exome sequencing (WES) release from the UKB, 49,960 samples had undergone exome capture using the IDT xGen Exome research panel v1.0. Samples were sequenced using the Illumina Novaseq 6000 platform and S2 flow cells with dual-indexed 75 × 75 paired-end reads (UKB, 2021). All common variants (MAF >= 0.01) were removed prior to analysis, and variant and sample QC was pre-performed as previously described (Van Hout *et al*., 2020). Briefly, any SNVs that did not have a read depth >=7 was changed to no-calls. SNVs which met at least one of the two following criteria were kept for further analysis: 1) at least one heterozygous variant genotype with an allele balance ratio greater or equal to 15%; 2) at least one homozygous variant genotype. For indels, variants with a minimum read depth of >=10 and an allele balance of >= 0.20 were kept. Additionally, variants with a sample-wise missingness of >=5% were also removed.

Samples which showed discordance between self-reported and genetically determined sex, high rates of heterozygosity, low overall exome sequence coverage (less than 85% coverage achieving 20x depth), and genetically identified duplicate samples were removed from the analysis. Since the WES dataset contains far fewer common variants used to perform QC than the genotyping array, we also chose to apply the quality control standards for PC-corrected heterozygosity, relatedness (relative cut-off >= 0.125), sex discordance, and sex chromosome aneuploidies as established by Bycroft *et al*., 2018. Specifically, in instances where two or more individuals were deemed related, one individual from the related group was randomly chosen to be kept for analysis, and the others removed. In total, a maximum of 39,274 individuals were included in the analysis.

### Phenotype definitions

#### PMMO

For the PMMO analyses, all baseline pre-surgery traits were recorded as close to the surgery date as possible. The median date of baseline measurements was 28.2 days prior to surgery. In total, four traits were analysed at baseline (weight, BMI, HbA1c, Type 2 diabetes status), and five traits at 1-year and 2-years after surgery (weight, BMI, % weight loss, % excess weight loss, and HbA1c).

#### UK biobank

Baseline measures of weight, waist circumference, hip circumference, body fat percentage (as determined by bioelectrical impedance), unfasted blood glucose, HbA1c, collected at recruitment, were used as the quantitative outcome variables for all UKB analyses. Obesity was defined as BMI >= 30 kg/m^2^. Waist-hip ratio was calculated using waist circumference (cm) / hip circumference (cm). Type 2 diabetes was defined in the UKB as having a previous record of “*diagnosis: ICD-10: E11 non-insulin dependent diabetes mellitus”* or a HbA1c > 48mmol/mol at the time of enrolment. Individuals with other types of diabetes (“*diagnosis summary - ICD-10: E10, E13, E14”*) or gestational diabetes (“*Medical conditions: gestational diabetes only”*) were removed from analyses. Individuals with operating procedure codes suggestive of bariatric surgery which could lead to weight loss were also removed (NHS digital, 2022). For blood glucose levels, only individuals who were noted to be fasted were included for analysis.

### Genetic analysis of rare *GLP1R* variants in the PMMO cohort

The minor allele frequencies (MAF) of the *GLP1R* variants genotyped in the PMMO cohort were compared to the overall frequencies of the same variants in the gnomAD V2.1 dataset (Karczewski *at al*., 2020). CADD PHRED scores (Kircher *et al*., 2014; Rentzsch *et al*., 2019) were used as a prediction of likely functional deleteriousness. American College for Medical Genetics (ACMG) scores were calculated using VarSome (Richards *et al*., 2015; Kopanos *et al*., 2019). Fisher’s exact test was used to assess whether *GLP1R* variants were over-represented in the PMMO cohort compared to the gnomAD controls dataset.

To determine the effect of *GLP1R* variants on baseline characteristics before bariatric surgery (weight, BMI, HbA1c, and T2D status), two-tailed one sample t-tests were used for continuous outcomes, and Fisher’s exact test for dichotomous outcomes. The same process was applied to the measurements at one and two years of follow up for both weight (weight, BMI, percentage weight loss, and percentage excess weight loss), and Type 2 diabetes-related outcomes (non-fasting blood glucose, HbA1c, and T2D status).

### The effect of rare *GLP1R* variants in the UK biobank

Only 4/8 *GLP1R* rare variants that we found in the PMMO study were also available in the UKB WES data in unrelated individuals of European ancestry. To determine whether these 4 *GLP1R* variants influenced the baseline weight-related- (waist circumference, hip circumference, weight, BMI, waist-hip ratio, obesity status, and body fat percentage) and T2D-related (blood glucose, HbA1c, and type 2 diabetes status) -phenotypes, two-tailed one sample t-tests for continuous variables and Fisher’s exact tests for dichotomous variables were used.

### The effect of rare GLP1R variants on weight and Type 2 diabetes-related traits in the UK biobank

To determine whether the *GLP1R* gene was associated with weight and glycaemic traits in the UKB, we performed collapsing gene-based testing. The population for this analysis was restricted to 39,274 unrelated individuals of European ancestry in the UKB, since multiple ancestry variance-based gene-based tests have yet to be developed. The WES data from the UKB was annotated with gene names using snpEff v5.1 and genome build version GRCh38. Only variants which were: 1) predicted to occur in the exonic or splice-site regions of *GLP1R* according to CCDS release 22, 2) were rare (MAF >= 0.01), or 3) were not exceedingly rare (minor allele count < 5; ∼ MAF >= 0.0002) in this cohort were omitted. Variants which had predicted pathogenicity (a CADD PHRED score of 15 or above) kept for analysis. Any frameshift, stop-gained, and essential splice variants that were deemed “low confidence” by LOFTEE (Karczewski *et al*., 2020) were also not included for analysis. In total, 17 rare variants were used in the burden testing, which included 2,513 individuals that carried at least one rare *GLP1R* variant and 36,761 controls who were homozygous for the major allele for all *GLP1R* mutations.

## Results

Summary statistics for the samples used in these analyses are available in supplementary Table 1. In total, 8 *GLP1R* rare variants were identified in 34/1714 (1.98%) PMMO participants (for comparison, the sum of MAFs for these 8 variants in gnomAD controls was 0.0067). The 8 variants had a mean CADD-PHRED score of 22.4 (Table 1).

**Table 1.** Rare and potentially deleterious *GLP1R* variants carried in the PMMO. POS, position; REF, reference allele; ALT, alternate allele; CADD, CADD phred score; ACMG, American College for Medical Genetics guidelines score; LB, likely benign; VUS, variant of unknown significance; LP, likely pathogenic; MAF, minor allele frequency. UKBB, UK Biobank POS refers to genome build GRCh38. NA represents unavailability of the variant in UK biobank dataset because the locus was not covered by the capture methodology for whole exome sequencing.

| RSID | POS | REF | ALT | CADD<br>PHRED | ACMG | N<br>PMMO | MAF<br>PMMO | MAF UKBB exome<br>sequence dataset | MAF gnomAD<br>controls | MAF in highest<br>ethnic<br>subpopulation<br>in gnomAD |
| --- | --- | --- | --- | --- | --- | --- | --- | --- | --- | --- |
| rs10305421 | 39048899 | G | A | 23.2 | LB | 16 | 0.00467 | 0.0056 | 0.0037 | 0.022 |
| rs866312541 | 39056410 | C | T | 25.4 | VUS/LP | 1 | 0.00029 | 0.000013 | 0 | 0 |
| rs1483005423 | 39073647 | A | G | 27.4 | VUS/LP | 1 | 0.00029 | NA | 0.0000090 | 0.0001105 |
| rs145619754 | 39073748 | C | T | 23.2 | LB | 6 | 0.00175 | 0.00020 | 0.00016 | 0.0004462 |
| rs202171972 | 39079189 | C | A | 24.6 | LB | 3 | 0.00088 | 0 | 0.00021 | 0.0014 |
| rs200065197 | 39079646 | C | T | 25.3 | LB | 1 | 0.00029 | NA | 0.000042 | 0.00024 |
| rs146868158 | 39085942 | C | T | 23.8 | LB | 1 | 0.00029 | 0.00030 | 0.0003742 | 0.00097 |
| rs201672448 | 39086015 | G | C | 17.8 | LB | 5 | 0.00146 | 0 | 0.002166 | 0.015 |

Considered individually, five of the eight variants appeared numerically over-represented in the PMMO cohort (see table 1), but this did not reach statistical significance compared to gnomAD controls. Although all variants were rare in the gnomAD controls overall, 2/8 (rs201672448, and rs10305421) were more common (MAF > 0.01) in at least one ethnic sub-population in the gnomAD data (Table 1). Of the 8 variant loci, 6 were covered by the UKB whole exome sequencing (WES) capture methodology, and only 4/6 of the analysable variants were observed in the UKB participants (Table 2). In total, there were 459 carriers of these 4 *GLP1R* variants, with allele frequencies that were comparable to gnomAD, but similar than in the PMMO cohort, for all variants (Supplementary Table 3).

**Table 2.** Summary statistics for carriers and non-carriers of rare and potentially deleterious *GLP1R* variants in the PMMO cohort for pre- and post-bariatric surgery. Values in brackets indicate standard deviation.

| Traits | GLP1R carrier status |  | Data availability (n) |  | P-value |
| --- | --- | --- | --- | --- | --- |
|  | Carriers | Non-carriers | Carriers | Non-carriers |  |
| **Pre-surgery** |  |  |  |  |  |
| Age (Years) | 46.5(11.7) | 46.05(11.3) | 33 | 1529 | 0.90 |
| Sex (Male, %) | 24.2 | 26.4 | 34 | 1600 | 0.92 |
| Ethnicity (European, %) | 58.8 | 62.1 | 34 | 1600 | 0.43 |
| weight (Kg) | 126.4 | 128.8 | 23 | 1128 | 0.66 |
| BMI (Kg/m2) | 45.0(7.9) | 46.3(8.38) | 21 | 931 | 0.66 |
| Type 2 Diabetes (%) | 37.5 | 31.5 | 32 | 1613 | 0.60 |
| HbA1c (mmol/mol) | 46.2(13.4) | 46.5(13.7) | 5 | 57 | 0.63 |
| underwent bariatric surgery (%) | 71.0 (n = 24) | 73.7 (n = 1239) | 34 | 1680 | 0.78 |
| Gastric bypass (% of bariatric surgeries) | 25.4 | 34.3 | 24 | 1239 | 0.14 |
| Gastric sleeve (% of bariatric surgeries) | 35.2 | 34.0 | 24 | 1239 | 0.92 |
| **1-year follow-up** |  |  |  |  |  |
| Weight (Kg) | 92.2(21.8) | 94.2 (30.6) | 24 | 1128 | 0.67 |
| BMI (Kg/m2) | 33.1(6.6) | 33.2(6.7) | 20 | 831 | 0.54 |
| weight loss (raw; Kg) | 33.6(15.8) | 34.9(16.4) | 15 | 390 | 0.79 |
| percentage total weight loss (%) | 25.9(8.5) | 26.4(10.5) | 14 | 233 | 0.47 |
| HbA1c (mmol/mol) | 41.7(12.3) | 38.5(9.85) | 11 | 588 | 0.41 |
| **2-year follow up** |  |  |  |  |  |
| Weight (Kg) | 91.8(13.7) | 96.4(23.8) | 18 | 557 | 0.77 |
| BMI (Kg/m2) | 34.5(14.7) | 34.3(7.6) | 17 | 548 | 0.32 |
| weight loss (raw; Kg) | 34.6(10.1) | 35.5(15.0) | 13 | 322 | 0.50 |
| percentage total weight loss (%) | 25.8(10.3) | 26.9(11.5) | 13 | 233 | 0.47 |
| HbA1c (mmol/mol) | 40.8(11.3) | 39.2(11.0) | 8 | 397 | 0.79 |

In the small group of PMMO carriers, no significant associations were found between the 8 *GLP1R* variants, and anthropometric or surgery-related phenotypes at baseline, or surgery outcomes. (Table 2). There was also no association of the 4/8 available variants with obesity-related phenotypes in the UKB dataset (Table 3).

**Table 3.** Phenotype summary statistics for the four rare and potentially deleterious *GLP1R* variants identified in the PMMO cohort in the UK biobank. Values in brackets represent standard deviation. P-values indicate 2-sided Student’s T-test for continuous outcomes, and Fisher’s exact test for dichotomous outcomes.

|  | Carriers | non-Carriers | P-value |
| --- | --- | --- | --- |
| Number | 629 | 37248 | NA |
| Age at recruitment (years) | 56.9(8.3) | 56.9(7.9) | 0.92 |
| Sex (% Male) | 46.1 | 46.3 | 0.30 |
| Weight (kg) | 77.8(14.8) | 78.4(15.8) | 0.43 |
| Body mass index (kg/m2) | 27.4(4.65) | 27.3(4.7) | 0.83 |
| Waist circumference (cm) | 94.4(12.4) | 90.1(13.4) | 0.24 |
| Hip circumference (cm) | 102.7(8.9) | 103.1(9.1) | 0.31 |
| Waist-hip ratio | 0.87(0.09) | 0.87(0.09) | 0.61 |
| Body fat percentage (%) | 32.0(8.7) | 31.4(8.5) | 0.19 |
| Obesity (%) | 24.2 | 23.7 | 0.83 |
| Severe obesity (%) | 2.0 | 1.7 | 0.72 |
| Type 2 diabetes status (%) | 9.2 | 6.3 | **0.02** |
| Blood glucose (mmol/mol) | 5.1(1.1) | 5.0(0.9) | 0.94 |
| HbA1c (mmol/mol) | 35.9(6.6) | 35.5(5.6) | 0.20 |

The small number of GLP1R variant carriers in the PMMO also limited our power for analysis of T2D status, but we noted that although non-statistically significant, T2D prevalence at baseline was higher in *GLP1R* variant carriers than in non-carriers (37.4% vs 31.5%, P = 0.60). Accordingly, we examined the effect of these variants on T2D in the UKB dataset: carriage of any of the 4/8 analysable *GLP1R* variants was significantly associated with increased likelihood of T2D (OR = 1.5(CI: 1.06-2.07), P = 0.015).

Since a statistically significant association with T2D was found using only the subset of *GLP1R* variants that we had originally identified in the PMMO cohort, we took advantage of the exome sequence data in the UKB to carry out gene-based testing for *GLP1R*. Seventeen *GLP1R* variants were included in the gene-based testing due to having a CADD-PHRED score of >15, and a “high confidence” flag in LOFTEE if they were frameshift, stop-gain, or splice site variants (Supplementary table 3). In total, there were 1614 carriers of these 17 variants in the UK biobank 50k WES data.

Using a more stringent definition of predicted pathogenicity with a threshold of CADD-PHRED>20, we identified a subset of 629 individuals carrying one of 12 variants (Supplementary Table 3). There was a statistically significant association between *GLP1R* variant carrier status and T2D status in both groups (p = 0.0016, Table 4). There was also an association with HbA1c levels in UKB participants who had no diagnosis of any type of diabetes (P = 0.039, table 4) We further investigated the role of obesity in *GLP1R*-related T2D risk: the effect of *GLP1R* variant carriage on T2D status was statistically significant only in individuals who had obesity (OR = 1.60 (CI: 1.21-2.09), p = 6.8×10-4 for CADD 15; OR =2.09 (CI: 1.42-3.04), p = 1×10-4 for CADD 20). There was no statistically significant association in individuals without obesity (OR = 1.10 (CI: 0.78-1.51), p = 0.55 for CADD 15; OR = 1.14(CI: 0.67-1.81), p =0.53 for CADD 20).

**Table 4.** Summary statistics for the 17 rare and potentially deleterious *GLP1R* variants included in the gene-based test in the UK Biobank for CADD 15 and CADD 20. Values in brackets represent standard deviation.

|  | CADD-PHRED 15 |  |  | CADD-PHRED 20 |  |  |
| --- | --- | --- | --- | --- | --- | --- |
|  | carriers | non-carriers | P-value | carriers | non-carriers | P-value |
| Number | 1614 | 35147 | NA | 629 | 36132 | NA |
| Age at recruitment (years) | 56.8(7.9) | 56.9(7.9) | 0.37 | 57.1(7.9) | 56.9(7.9) | 0.80 |
| Sex (% Male) | 46.1 | 43.1 | 0.030 | 46.0 | 44.2 | 0.35 |
| Weight (kg) | 77.6(15.2) | 78.4(15.8) | 0.07 | 77.9(12.8) | 78.4(0.42) | 0.41 |
| Body mass index (kg/m2) | 27.3(4.7) | 27.3(4.7) | 0.73 | 27.3(4.7) | 27.3(4.7) | 0.96 |
| Waist circumference (cm) | 89.5(13.0) | 90.1(13.4) | 0.08 | 89.5(12.8) | 90.1(13.4) | 0.25 |
| Hip circumference (cm) | 102.9(9.0) | 103.1(9.1) | 0.39 | 102.6(9.2) | 103.1(9.1) | 0.13 |
| Waist-hip ratio | 0.87(0.09) | 0.89(0.09) | 0.10 | 0.87(0.09) | 0.87(0.09) | 0.94 |
| Body fat percentage (%) | 31.9(8.5) | 31.4(8.5) | 0.096 | 31.8(8.7) | 31.5(8.3) | 0.35 |
| Obesity (%) | 23.7 | 23.8 | 0.95 | 23.7 | 23.8 | 0.96 |
| Severe obesity (%) | 1.75 | 1.84 | 0.76 | 1.74 | 2.07 | 0.54 |
| Type 2 diabetes status (%) | 8.2 | 6.2 | **0.004** | 9.5 | 6.2 | **0.0016** |
| Blood glucose (mmol/mol) | 5.0(0.8) | 5.1(0.8) | 0.51 | 5.1(1.12) | 5.1(0.91) | 0.71 |
| HbA1c (mmol/mol) | 35.9(6.0) | 35.6(5.6) | **0.028** | 36.2(7.3) | 35.6(5.7) | **0.039** |

## Discussion

We used a custom AXIOM genotyping array to screen 1714 unrelated adults with severe obesity from the PMMO cohort and identified 34 carriers of 8 rare and potentially deleterious *GLP1R* variants. Subsequent analysis of the 4 of the 8 variants available in the UKB cohort 50K WES dataset revealed a significant association with T2D. This prompted a gene-level analysis of rare, predicted-deleterious (according to CAD-PHRED score >15, and >20) *GLP1R* variants in the exome sequencing data from the UKB which confirmed the association with T2D, and revealed an association with HbA1c levels in UKB participants.

Although there was no evidence of association with obesity *per se*, (either in PMMO or the UKB), when stratified by obesity, Carriage of a variant with CADD-PHRED score>20 doubled the risk of T2D in UKB participants with BMI >30 kg/m^2^. GLP1 and GLP1R are important regulators of post-prandial glycaemic control (as reviewed by Hinnen *et al*., 2017), probably explaining the observation that rare and potentially deleterious *GLP1R* variants are associated with T2D status at recruitment in the UK biobank. The observed odds ratios for *GLP1R* rare variant carriage represent the composite effect of all variants analysed: there may be some specific variants that decrease risk of T2D, as well as others that increase it. Our analysis included the analysis of data from the 50K release of UKB exome sequencing data, but larger studies, as well as multi-ethnic analyses, may be required to ascertain the functional implications of each individual variant. Nonetheless, the composite effect exceeds that of rs790314; the common *TCF7L2* variant with largest effect size identified by the current largest GWAS of T2D risk (OR = 1.38) and is comparable to the effect other rare T2D susceptibility variants such as rs536643418 and rs140242150 in TCF7L2 (ORs 1.65 and 1.39, respectively) and rs571342427 in INS/IGF2 (OR = 1.89) (Mahajan *et al*., 2018). It remains possible that some specific *GLP1R* variants may confer higher risk whilst others are protective (as observed by rare *MC4R* variants, some of which increase the risk of obesity, whilst others protect against it (Lotta *et al*., 2019)).

A major challenge in this work is the limited statistical power afforded by the limited sample sizes with robust rare variant data available. To fully understand the phenotypic implications of individual rare variants, particularly variants of unknown significance (VUS), ultra-large-scale collaborations between investigators are essential. Consortia, such as the Biobank Rare Variant Analysis (BRaVa) consortium, have recently formed in attempt to address such issues.

The obesity-specific effect of *GLP1R* variants on T2D risk in the UK biobank also requires further investigation. In general, although some GLP1 agonists are very effective in treatment of obesity, we saw no association with obesity or any of the adiposity-related outcomes we examined in PMMO or UKB. The PMMO cohort, although sizable for a clinical cohort with an extreme phenotype, remains very small for rare variant analysis, especially where other major effects of variants in other genes may also be operating. Severe obesity is a complex phenotype: with 5-10% of individuals with a BMI >35 kg/m^2^ reported to carry monogenic mutations (Lin *et al*., 2021; Loos *et al*., 2022). Whilst data exists for the most well-recognised “obesity genes”, in general the implications of rare variants of other genes in the context of severe obesity are poorly understood. There was also low power for analysis of participants with a BMI >30 kg/m^2^ in the UKB dataset since it exhibits health volunteer bias. Whilst the UKB provides an excellent resource for individuals with healthy BMI, only ∼800 individuals included in the 50k exome release data have severe obesity, and of these, 302 UKB participants who had undergone bariatric surgery had to be excluded from the analysis, since pre- and post-surgery obesity-related phenotypes are unavailable.

Compounding the above challenges, our PMMO analysis is likely to be missing some *GLP1R* variants. For reasons of cost and efficiency, a custom genotyping array was utilised in analysis of the PMMO cohort, meaning that only variants included in the array design could be tested (117 variants, as opposed to the 478 variants observed in the UK biobank 50k exome data release interrogated here), so any other *GLP1R* variants will have been missed. This means that our finding of a prevalence of around 2% *GLP-1R* variant carriers in PMMO is likely to be an underestimate. Exome sequencing of this cohort may provide further information.

GLP1 agonists are now widely used in the treatment of obesity and diabetes and there is an obvious question about how genetic variants in the GLP1R receptor affect the efficacy of these agents. Large-scale analysis of clinical data will be necessary to explore this, which may well require global collaboration. In the same way, analysis of much larger cohorts of patients undergoing bariatric surgery will be necessary to explore implications of GLP1R variants for surgery outcomes, including both weight loss and remission of T2D.

In summary, although our work has unavoidable power limitations, we provide compelling evidence that rare deleterious *GLP1R* variants increase the risk of T2D risk in individuals within the UK biobank, and particularly in individuals with obesity. Given these data, future studies should focus on determining whether *GLP1R* variants can help us understand the variation in outcomes following therapeutic administration of GLP1R agonists and should utilise larger sample sizes to re-affirm the findings of this research in the context of more severe forms of obesity.

## Data Availability

All data produced in the present study are available upon reasonable request to the authors.

